# Prevalence of and risk factors for Tuberculosis among health care workers in Yogyakarta, Indonesia

**DOI:** 10.1101/2022.12.04.22283080

**Authors:** Stephanie Main, Rina Triasih, Jane Greig, Arif Hidayat, Immanuel Billy Brilliandi, Syarifah Khodijah, Geoff Chan, Nova Wilks, Amy Elizabeth Parry, Betty Nababan, Philipp du Cros, Bintari Dwihardiani

## Abstract

**Background:** Healthcare workers (HCWs) are at risk of contracting TB, particularly when in high tuberculosis (TB) burden settings. Routine surveillance data and evidence are limited on the burden of TB amongst HCWs in Indonesia.

**Objective:** To measure the prevalence of TB infection (TBI) and disease among HCWs in four healthcare facilities in Yogyakarta and explore risk factors for TBI.

**Methods:** A cross-sectional TB screening study targeted all HCWs from four pre-selected facilities (1 hospital, 3 primary care) in Yogyakarta, Indonesia. Voluntary screening included symptom assessment, Chest X-ray (CXR), Xpert MTB/RIF (if indicated) and tuberculin skin test (TST). Analyses were descriptive and included multivariable logistic regression.

**Results:** Of 792 HCWs, 681 consented (86%) to the screening; 59% (n=401) were female, 62% were medical staff (n=421), 77% worked in the one participating hospital (n=524), and the median time working in the health sector was 13 years (IQR: 6-25 years). Nearly half had provided services for people with TB (46%, n=316) and 9% reported ever having TB (n=60).

Among participants with presumptive TB (15%, n=99/662), none were diagnosed microbiologically or clinically with active TB disease. TBI was detected in 25% (95% CI: 22-30; n=112/441) of eligible HCWs with a TST result. A significant association was found between TB infection and being male (adjusted Odds Ratio (aOR) 2.02 (95%CI: 1.29-3.17)), currently working in the participating hospital compared to primary care (aOR 3.15 (95%CI: 1.75-5.66)), and older age (1.05 OR increase per year of life between 19-73 years (95%CI: 1.02-1.06)).

**Conclusion:** This study supports prioritisation of HCWs as a high-risk group for TB infection and disease, and the need for comprehensive prevention and control programs in Indonesia. Further, it identifies characteristics of HCWs in Yogyakarta at higher risk of TBI, who could be prioritised in screening programs if universal coverage of prevention and control measures cannot be achieved.

## INTRODUCTION

Tuberculosis (TB) remains one of the most burdensome and deadliest infectious diseases worldwide, causing an estimated 10.6 million cases and 1.6 million deaths in 2021 (1). Low- and middle-income countries (LMICs) bear the brunt of TB’s impact, with more than 95% of deaths occurring in these countries and only eight LMICs accounting for two thirds of the global caseload (1).

Healthcare workers (HCWs), are at an increased risk of contracting TB compared to the general population (2–4). TB infection (TBI) risk is higher in LMICs where exposure to infected individuals is more likely, and infection prevention and control practices might be insufficient or inconsistent (5). TB burden estimates among HCWs in LMICs vary considerably. A systematic review published in 2019 found TBI prevalence by positive tuberculin skin test (TST) ranged from 14–98% (mean 49%) among HCWs in LMICs, with higher prevalence among HCW populations in higher TB burden settings (defined as ≥300 cases per 100,000 population)(6). Earlier reviews reported similar variability with annual TBI incidence among HCWs ranging between 0.5-14.3% (5,7).

Meeting ambitious End TB strategy targets by 2035 requires a well-resourced, skilled and healthy healthcare workforce (8). The burden of TB in HCWs highlights the negative impact occupational TB and nosocomial TB transmission could have on meeting End TB targets and wider TB elimination. The burden also highlights the need for further research, and targeted policy and programming among this high priority population.

Indonesia has the third highest TB caseload internationally and an annual disease incidence of 301 per 100,000 (9). Surveillance data on the burden of TB among HCWs in Indonesia are limited, with estimates of reported TBI burden ranging from 24% among HCWs in primary healthcare centres (PHCs) to 77% in tertiary hospitals (10,11). There is limited information on TB burden in HCWs across many provinces including Yogyakarta. We aimed to measure the prevalence of infection and disease among HCWs in four healthcare facilities in Yogyakarta and explore risk factors for infection.

## STUDY POPULATION, DESIGN AND METHODS

### Study setting

Yogyakarta province, located in central southern Java, has an estimated TB disease incidence rate between 250-300 per 100,000 population and an estimated case detection rate of 34.2% (12). The Yogyakarta health system, like Indonesia’s, encompasses several jurisdictional levels and includes both public and private providers (13). District level public and private hospitals along with PHCs provide TB services comprising case detection, diagnosis, treatment and prevention (13).

### Study design, site selection and study population

The study selected all healthcare facilities delivering TB care within two sub-districts of two districts of Yogyakarta province. Selection of healthcare facilities in this study was targeted, based on the respective sub-district’s involvement in a TB Reach funded active case finding program (14). Health office consultation, low case detection rates, geography and operational challenges such as near absence of contact tracing and testing, were reasons for sub-district selection into the program. The healthcare facilities selected consisted of one private hospital treating between 100-120 TB patients per year (2018-2019) and three PHCs treating 4-20 TB patients per year (2018-2019).

The target population for this cross-sectional study was all healthcare facility staff from the four pre-selected facilities aged ≥17 years, including non-medical (e.g., management and support services) and medical staff. There were 792 HCWs across the four facilities; 613 at the private hospital and 179 staff across the PHCs.

### Participant recruitment

Information about the screeening, including its purpose, date, and location were disseminated through advertisements in staff break rooms, online notification through the facility’s human resource network, and awareness-raising sessions for staff before the period of screening. The sessions were conducted by the study team and included information on TB, TB burden in HCWs, the value of screening, the screening process, dates and location of the screening, the consent process and participant confidentiality. The study was voluntary, free of charge, and participants did not receive incentives for participating.

### Data collection

Screening of health workers was conducted for one week at each PHC and two weeks at the private hospital between December 2020 to May 2021. HCWs were invited to join the study at a time that was convenient to them, which could be prior to, during or after work hours. TB infection and disease screening included symptom assessment, Chest X-ray (CXR), one spot sputum sample for GeneXpert MTB/RIF testing (if indicated) and TST. TBI was classified as TST positivity (>10mm induration or >5mm in immune suppressed participants) with no evidence of clinically manifest active TB (15,16). Standard COVID-19 infection prevention and control procedures and strategies were implemented throughout the study, with participants screening positive for COVID-19 symptoms offered a PCR test. People who tested positive for COVID-19 and/or those who had recently received a COVID-19 vaccine were followed up for TB screening during additional screening days. All tests were performed by trained healthcare professionals. Final TB diagnoses were made by the study team doctor, based on Indonesian and WHO definitions/criteria. Results and information on treatment options were disseminated by formal letter if the participant screening outcome was negative, or through a doctor consultation if TBI was diagnosed. Pending consent, participant eligibility to take TB preventive treatment (TPT) was assessed, and when relevant participants were supported to attend their chosen healthcare facility to initiate TPT according to the National guidelines.

Interview and examination data were entered electronically at the point of care using REDCap (17). The study interview collected data on participant demographic, clinical and occupational information including age, sex, healthcare role, time working in the health sector, history of TB, co-morbidities and other risk factors. All information was voluntary to disclose.

### Data Analysis

Re-identifiable study data were exported into Stata v15 (StataCorp, College Station, TX, USA). Descriptive analyses were utilised to measure TBI prevalence and examine cohort characteristics. Categorical variables were reported as numbers and proportions. Continuous variables were assessed for normality and reported as median and interquartile range (IQR).

Risk factors associated with TBI were investigated by examining demographic, occupational, and clinical factors using univariable and multivariable logistic regression. The effect was expressed as an unadjusted and adjusted odds ratio (OR) with 95% confidence intervals (95% CI). Variables were added to the multivariable model if p < 0.2 in the univariable analyses. Those recorded only for sub-sets of the sample or demonstrating collinearity were excluded. Backward selection was employed, and selection stopped when the multivariable model Likelihood Ratio test showed only a statistically non-significant improvement in modelling. Levels of significance in the multivariable model and statistical tests were set at 5%.

### Ethical Approval

Ethical approval was obtained from the Institutional Ethical Review Board at the University of Gadjah Mada, Indonesia and the Alfred Hospital Human Research Ethics Committee, Australia.

## RESULTS

Participation in the study was high with 86% (n=681/792) of registered HCWs at the facilities participating, ranging from 73-94% by facility. Of the 681 participants the median age was 40 years (IQR:30-48), 59% (n=401) were female, 71% (n=486) reported completing a university degree, and participant’s median household size was 4 people (IQR:2-5 people) (Table 1). Nearly two thirds of participants identified as medical staff (62%, n=421), mainly nurses (n=250, 37%), 77% worked in the hospital (n=524), and the median time working in the health sector was 13 years (IQR: 6-25 years) (Table 1). Nearly half had provided services for people with TB (46%, n=316) and 36% (n=243) had received TB training outside of their professional qualification.

**Table 1.** Demographic, occupational and clinical characteristics of healthcare workers screened for TB in primary health facilities and a private hospital in two sub-districts of Yogyakarta Indonesia

| Characteristics | Total (n,%) |
| --- | --- |
| **Total** | **681** |
| *Demographic Characteristics* |  |
| **Age** median [IQR] | 40 [30-48] |
| **Sex** |  |
| Male | 280 (41.1) |
| Female | 401 (58.9) |
| **Education** |  |
| High School Degree or Less | 195 (28.6) |
| University Degree | 486 (71.4) |
| **Household size**, median [IQR] | 4 [3-5]* |
| **Household expenses** |  |
| < Rp. 2.2 million per month (low) | 188 (27.6) |
| Rp. 2.2 - 5 million per month (medium) | 364 (53.5) |
| > Rp. 5 million per month (high) | 105 (15.4) |
| Missing | 24 (3.5) |
| *Occupational Characteristics* |  |
| **Health facility type** |  |
| Hospital | 524 (76.9) |
| Primary Health Centre | 157 (23.1) |
| **Occupation category** |  |
| *Medical* | 421 (61.8) |
| Nurse | 250 (37.0) |
| Dentist | 2 (0.3) |
| Laboratory Scientists | 18 (2.6) |
| Nutritionist | 15 (2.2) |
| Environmental Health | 2 (0.3) |
| Epidemiologist | 6 (0.9) |
| Radiographer | 9 (1.3) |
| Midwife | 47 (6.9) |
| Rehabilitation staff (e.g. physiotherapist) | 14 (2.1) |

| **Characteristics** | **Total**<br>**(n,%)** |
| --- | --- |
| Health Promotion Nurse | 2 (0.3) |
| Pharmacist and assistants | 43 (6.3) |
| Doctor | 12 (1.8) |
| Psychologist | 1 (0.1) |
| *Non-Medical* | **258 (37.9)** |
| Administrative staff | 82 (12.0) |
| Management | 4 (0.6) |
| Community Health Workers | 23 (3.4) |
| Other non-medical personnel (e.g. cleaners and cooks) | 149 (21.9) |
| *Missing* | **2 (0.3)** |
| **Length of time working in the health sector (years), median [IQR]** | **13 [6-25]** |
| **Length of time at this health facility (years), median [IQR]** | **11 [4-24]^** |
| **Provided services / care to a person diagnosed with tuberculosis** |  |
| No | 365 (53.6) |
| Yes | 316 (46.4) |
| **Length of time provided services / care to people with TB (years),**<br>**median [IQR]** | **7 [2-17]** |
| **Received TB training outside of professional qualification** |  |
| No | 436 (64.0) |
| Yes | 243 (35.7) |
| Missing | 2 (0.3) |
| **Received training on personal protective equipment** |  |
| No | 114 (16.7) |
| Yes | 565 (83.0) |
| Missing | 2 (0.3) |
| *Clinical characteristics* |  |
| **Nutritional status<sup>#</sup>** |  |
| Underweight (BMI < 18.5) | 41 (6.0) |
| Normal weight (BMI >= 18.5 to < 23) | 190 (27.9) |
| Overweight (BMI =>23 to < 25) | 126 (18.5) |
| Obese (BMI >= 25) | 324 (47.6) |
| **Smoking status** |  |
| No, never smoked | 488 (71.7) |
| No, but previously smoked | 94 (13.8) |
| Currently smoke | 99 (14.5) |
| **Self-reported diabetes mellitus status** |  |
| No | 617 (90.6) |
| Yes | 24 (3.5) |
| Unsure | 40 (5.9) |
| **Self-reported HIV Status** |  |
| Positive | 0 (0) |
| Negative | 190 (27.9) |
| Unsure | 491 (72.1) |
| **Self-reported Pregnancy Status Among Females** |  |
| No | 390 (97.3) |
| Yes | 11 (2.7) |
| **Lived with someone who had TB for at least one day** |  |
| No | 582 (85.5%) |
| Yes | 91 (13.4%) |
| Unsure | 8 (1.2%) |
| **Timing of living with someone with TB** |  |
| Before person with TB started treatment | 79 (86.8) |
| After person with TB started treatment | 11 (12.1) |
| Unsure | 1 (1.1) |
| **Previously been treated for TB** |  |
| No | 621 (91.2) |
| Yes, finished treatment | 59 (8.7) |
| Yes, currently on medication | 1 (0.1) |
| *Treated* |  |

\*656/681 participants responded
^ 678/681 participants responded

Among the 681 participants, 67% (n=450) were overweight or obese, 29% currently (15%, n=99) or previously (14%, n=94) smoked, self-reported diabetes mellitus (DM) prevalence was 4% (n=24), HIV status was known or disclosed among 28% (n=190) of participants and all were negative (18) (Table 1). Ninety-one participants (13%, n=91) reported ever having lived with someone who had TB, mostly (87%, n=79/91) before the person started TB treatment. Sixty participants reported ever having TB (9%, n=60), of whom 75% were females (n=45), 73% (n=44) were medical staff, the median time in the health sector was 10 years (IQR: 5-16 years), 60% (n=36) had provided services for people with TB, and 17% (n=10) had lived with someone with TB for at least one day (Table 2).

**Table 2:**
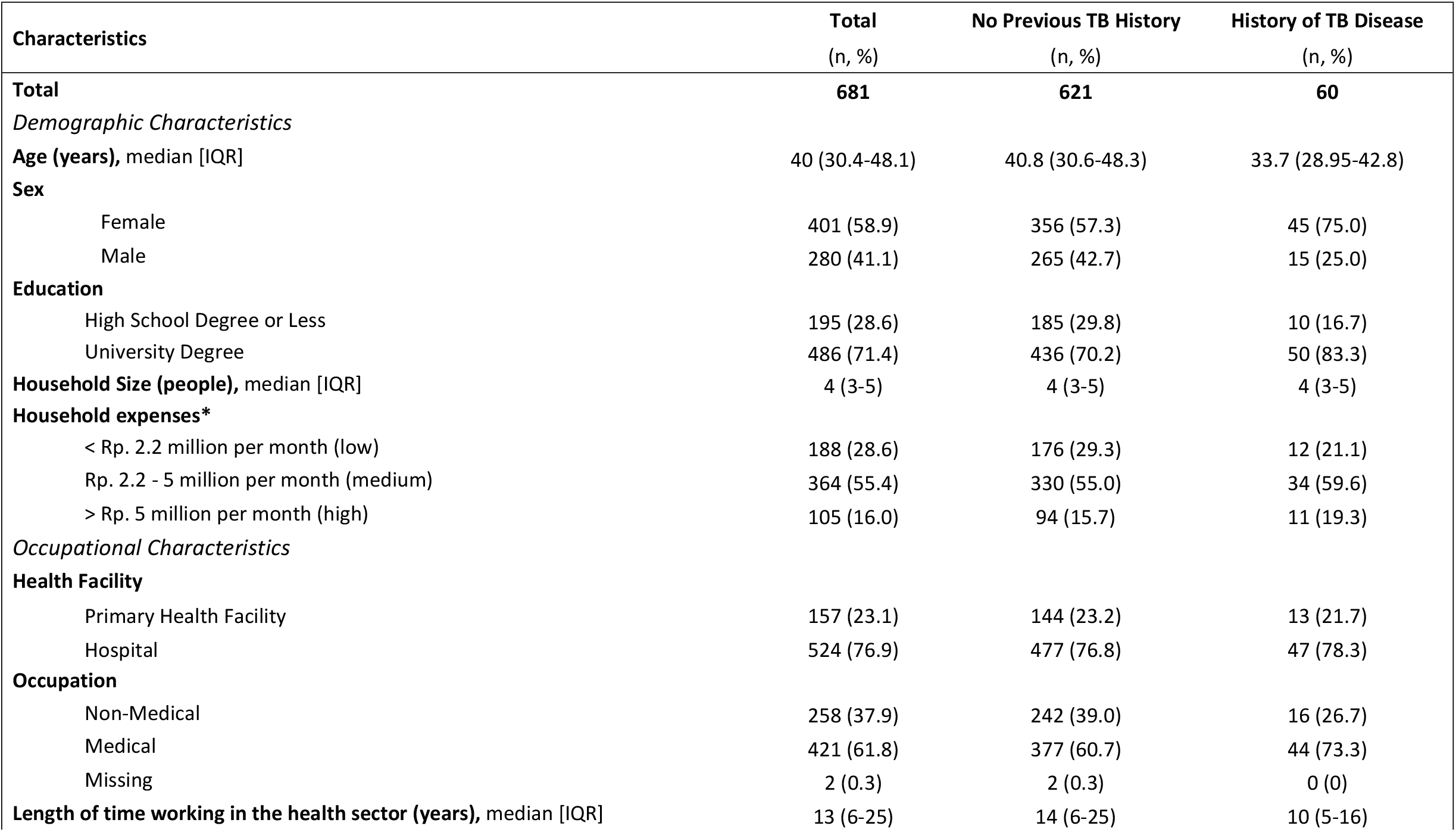
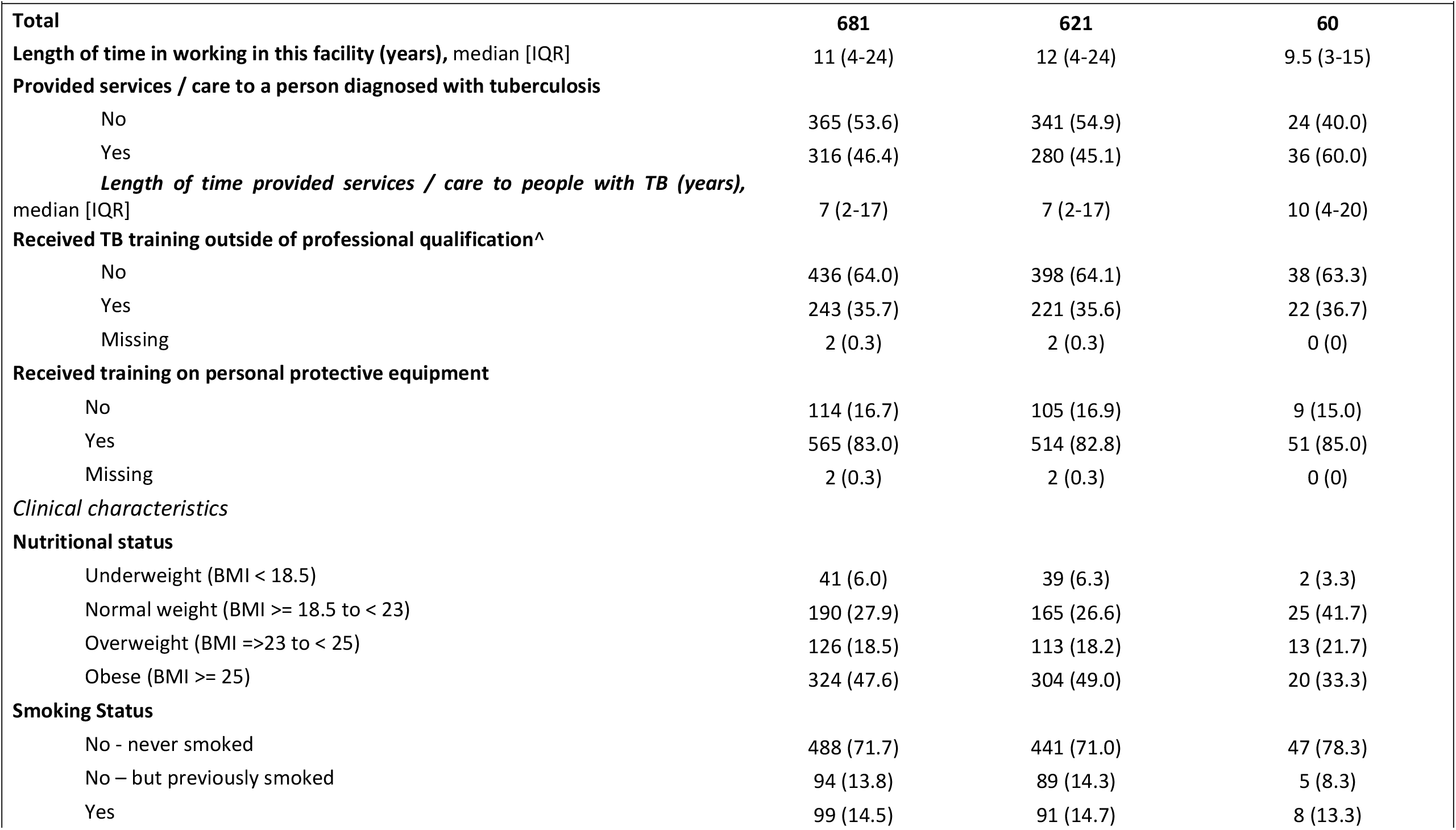
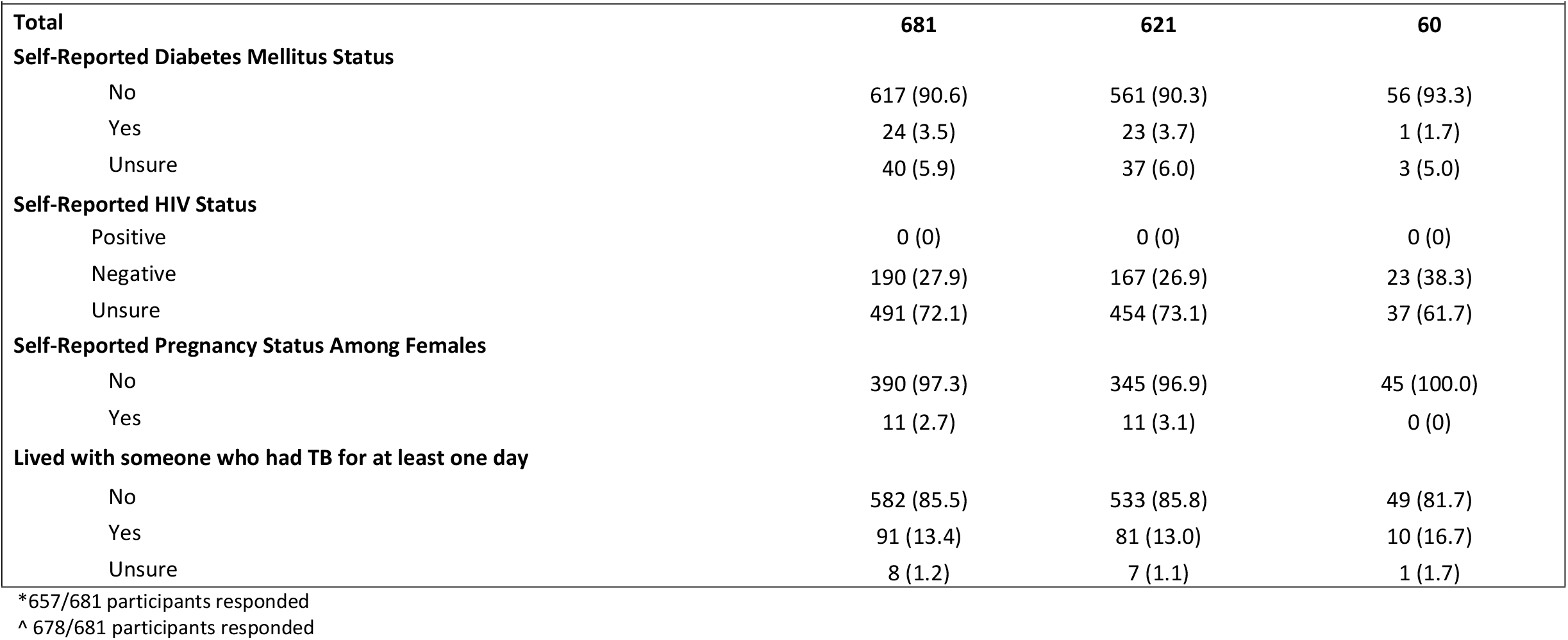
Characteristics associated with self-reported previous TB disease among healthcare workers in primary health facilities and a private hospital in two sub-districts of Yogyakarta Indonesia

During screening, 1% (n=5/681) of participants reported cough for more than two weeks or haemoptysis. Of those CXR screened (n=662; eight participants refused and 11 were pregnant), 36% had a lung or pleural abnormality (n=239/662), and 15% (n=97/662) of findings were consistent with possible active TB disease requiring further investigation and assessment (Table 3). Of the 15% of participants (n=99/662) with a combined symptom screen positive and/or a CXR suggestive of TB, none were diagnosed microbiologically (via Xpert MTB/RIF of sputum) or clinically with TB disease.

**Table 3:** Chest X-ray findings among healthcare workers with an abnormal chest X-ray in primary health facilities and a private hospital in two sub-districts of Yogyakarta Indonesia

| Chest X-Ray Abnormality | Suggestive of TB | Not Suggestive of TB | Total |
| --- | --- | --- | --- |
| **Total** | **N=97\*** | **N=147\*** | **N=239\*** |
| Infiltrates | 93 | 0 | 93 |
| Consolidation | 0 | 0 | 0 |
| Cavity | 1 | 0 | 1 |
| Calcification | 1 | 1 | 2 |
| Pleural Effusion | 2 | 0 | 0 |
| Fibrosis | 5 | 11 | 16 |
| Increased Bronchovascular Marking | 46 | 102 | 148 |
| Emphysematous Lung | 1 | 3 | 4 |
| Lymphadenopathy | 1 | 1 | 2 |
| Tented Diaphragm | 1 | 1 | 2 |

\*The denominator for the columns are the totals listed, not the total abnormalities listed. Some participants had more than one x-ray abnormality.

TST was offered to all participants who had not reported previously receiving TB treatment (n=621). It was administered to 78% (n=489/621) of those eligible and read in 90% (n=441/489) of them (Figure 1). TBI was diagnosed in 25% (95% CI: 22-30) of HCWs (n=112/441), most being HCWs in the hospital setting (86%; n=96/112) (Table 3). TBI prevalence differed between the hospital setting (31%, 95% CI: 26-37%; n=96/306) and PHCs (12%, 95% CI: 1-19%; n=16/135).

Of the 112 participants diagnosed with TBI, 8% (n=9) were underweight, 35% currently (19%, n=21) or previously (16%, n=18) smoked, 5% (n=6) reported having DM and 80% (n=90) were unsure of or unwilling to disclose their HIV status (Table 4). A significant association on multivariable logistic regression was found between TBI and being male (adjusted OR 2.02 (95%CI: 1.29-3.17)), currently working in the participating hospital compared to the PHCs (adjusted OR 3.15 (95%CI: 1.75-5.66)), and older age (1.05 OR increase per year of life between 19-73 years (95%CI: 1.02-1.06)) (Table 4).

**Table 4:**
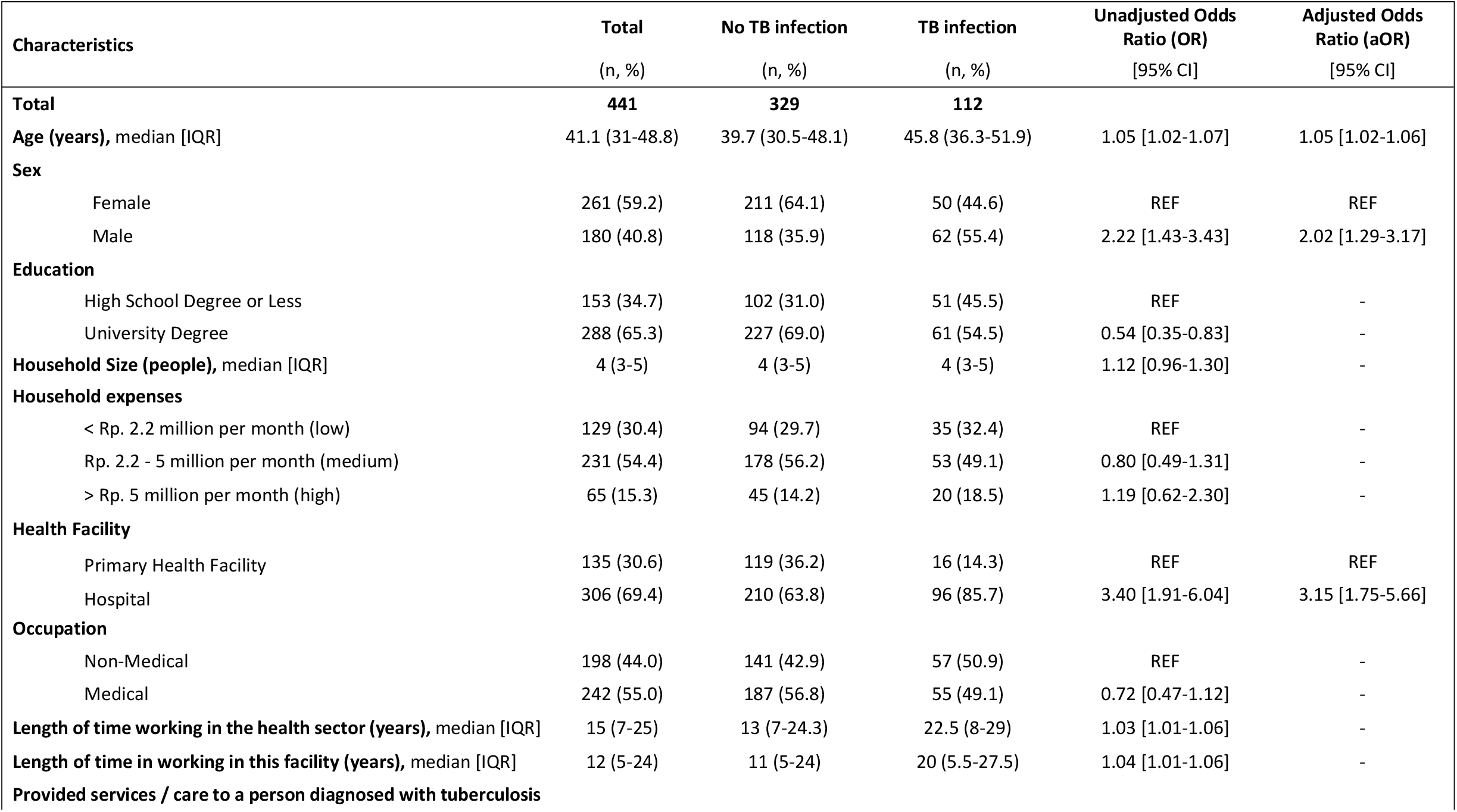
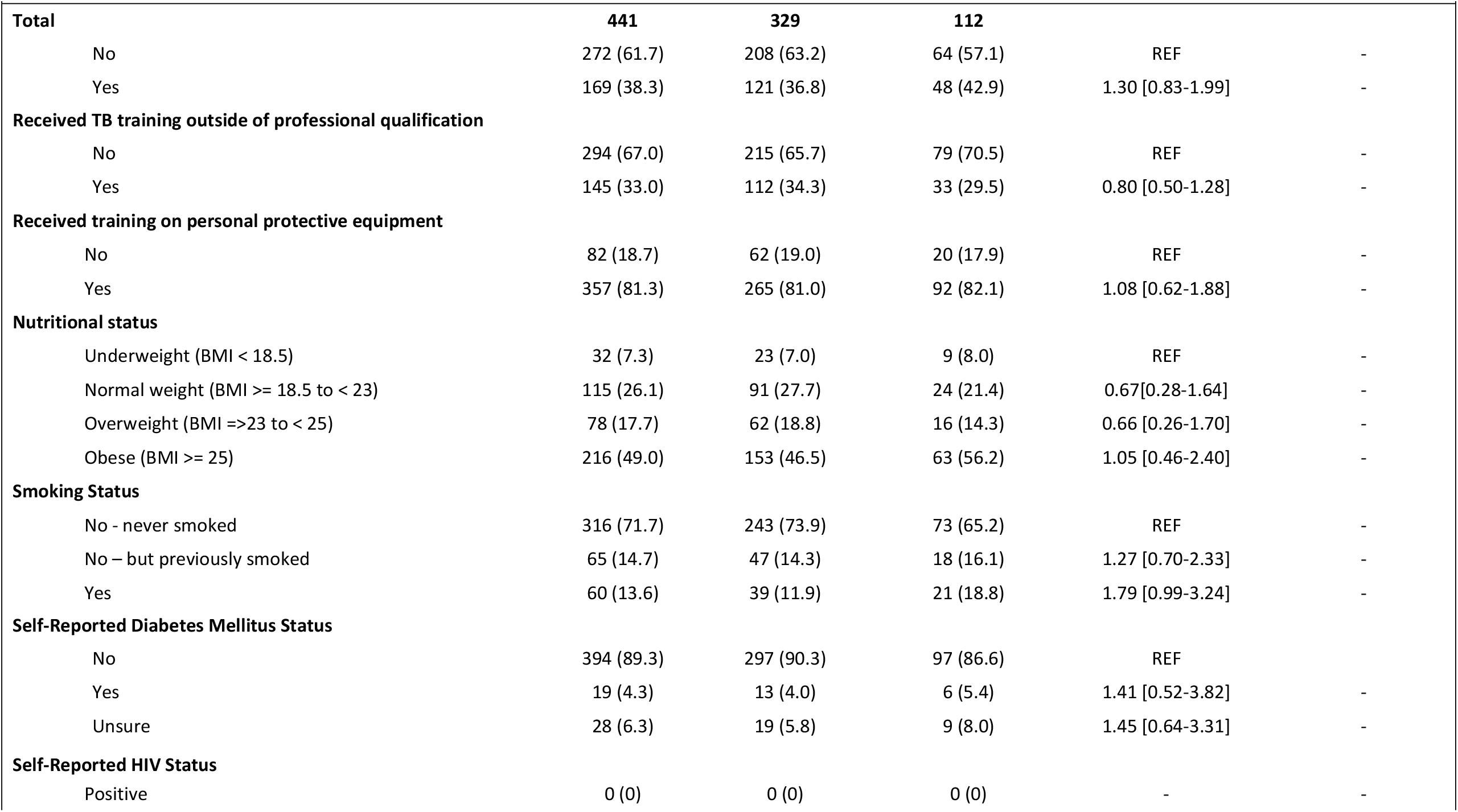
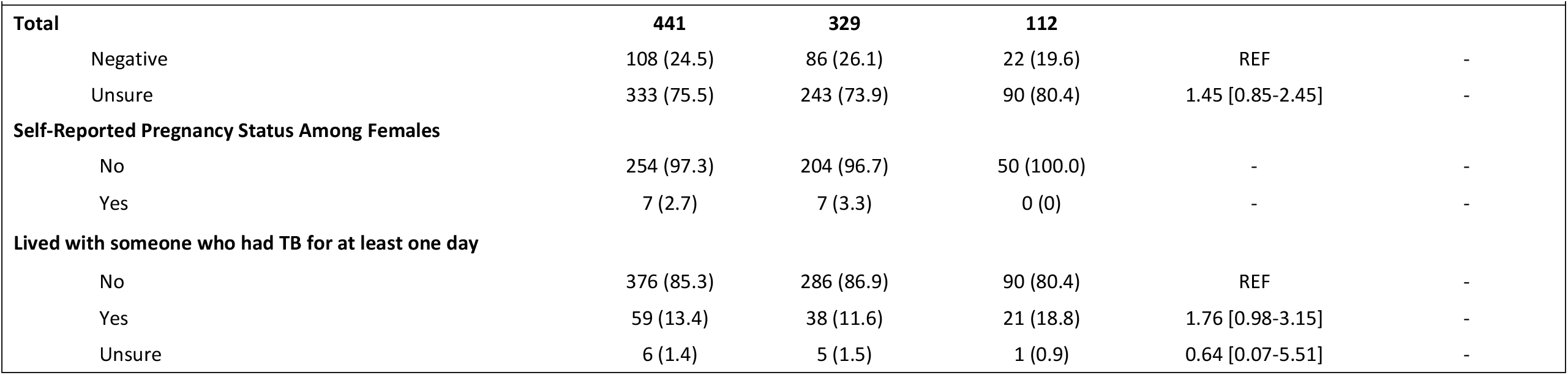
Characteristics associated with TB infection among healthcare workers in primary health facilities and a private hospital in two sub-districts of Yogyakarta Indonesia.

| Characteristics | Total<br>(n, %) | No TB infection<br>(n, %) | TB infection<br>(n, %) | Unadjusted Odds<br>Ratio (OR)<br>[95% CI] | Adjusted Odds<br>Ratio (aOR)<br>[95% CI] |
| --- | --- | --- | --- | --- | --- |
| **Total** | **441** | **329** | **112** |  |  |
| **Age (years), median [IQR]** | 41.1 (31-48.8) | 39.7 (30.5-48.1) | 45.8 (36.3-51.9) | 1.05 [1.02-1.07] | 1.05 [1.02-1.06] |
| **Sex** |  |  |  |  |  |
| Female | 261 (59.2) | 211 (64.1) | 50 (44.6) | REF | REF |
| Male | 180 (40.8) | 118 (35.9) | 62 (55.4) | 2.22 [1.43-3.43] | 2.02 [1.29-3.17] |
| **Education** |  |  |  |  |  |
| High School Degree or Less | 153 (34.7) | 102 (31.0) | 51 (45.5) | REF | - |
| University Degree | 288 (65.3) | 227 (69.0) | 61 (54.5) | 0.54 [0.35-0.83] | - |
| **Household Size (people), median [IQR]** | 4 (3-5) | 4 (3-5) | 4 (3-5) | 1.12 [0.96-1.30] | - |
| **Household expenses** |  |  |  |  |  |
| < Rp. 2.2 million per month (low) | 129 (30.4) | 94 (29.7) | 35 (32.4) | REF | - |
| Rp. 2.2 - 5 million per month (medium) | 231 (54.4) | 178 (56.2) | 53 (49.1) | 0.80 [0.49-1.31] | - |
| > Rp. 5 million per month (high) | 65 (15.3) | 45 (14.2) | 20 (18.5) | 1.19 [0.62-2.30] | - |
| **Health Facility** |  |  |  |  |  |
| Primary Health Facility | 135 (30.6) | 119 (36.2) | 16 (14.3) | REF | REF |
| Hospital | 306 (69.4) | 210 (63.8) | 96 (85.7) | 3.40 [1.91-6.04] | 3.15 [1.75-5.66] |
| **Occupation** |  |  |  |  |  |
| Non-Medical | 198 (44.0) | 141 (42.9) | 57 (50.9) | REF | - |
| Medical | 242 (55.0) | 187 (56.8) | 55 (49.1) | 0.72 [0.47-1.12] | - |
| **Length of time working in the health sector (years), median [IQR]** | 15 (7-25) | 13 (7-24.3) | 22.5 (8-29) | 1.03 [1.01-1.06] | - |
| **Length of time in working in this facility (years), median [IQR]** | 12 (5-24) | 11 (5-24) | 20 (5.5-27.5) | 1.04 [1.01-1.06] | - |
| **Provided services / care to a person diagnosed with tuberculosis** |  |  |  |  |  |
| **Total** | **441** | **329** | **112** |  |  |
| No | 272 (61.7) | 208 (63.2) | 64 (57.1) | REF | - |
| Yes | 169 (38.3) | 121 (36.8) | 48 (42.9) | 1.30 [0.83-1.99] | - |
| **Received TB training outside of professional qualification** |  |  |  |  |  |
| No | 294 (67.0) | 215 (65.7) | 79 (70.5) | REF | - |
| Yes | 145 (33.0) | 112 (34.3) | 33 (29.5) | 0.80 [0.50-1.28] | - |
| **Received training on personal protective equipment** |  |  |  |  |  |
| No | 82 (18.7) | 62 (19.0) | 20 (17.9) | REF | - |
| Yes | 357 (81.3) | 265 (81.0) | 92 (82.1) | 1.08 [0.62-1.88] | - |
| **Nutritional status** |  |  |  |  |  |
| Underweight (BMI < 18.5) | 32 (7.3) | 23 (7.0) | 9 (8.0) | REF | - |
| Normal weight (BMI >= 18.5 to < 23) | 115 (26.1) | 91 (27.7) | 24 (21.4) | 0.67[0.28-1.64] | - |
| Overweight (BMI =>23 to < 25) | 78 (17.7) | 62 (18.8) | 16 (14.3) | 0.66 [0.26-1.70] | - |
| Obese (BMI >= 25) | 216 (49.0) | 153 (46.5) | 63 (56.2) | 1.05 [0.46-2.40] | - |
| **Smoking Status** |  |  |  |  |  |
| No - never smoked | 316 (71.7) | 243 (73.9) | 73 (65.2) | REF | - |
| No – but previously smoked | 65 (14.7) | 47 (14.3) | 18 (16.1) | 1.27 [0.70-2.33] | - |
| Yes | 60 (13.6) | 39 (11.9) | 21 (18.8) | 1.79 [0.99-3.24] | - |
| **Self-Reported Diabetes Mellitus Status** |  |  |  |  |  |
| No | 394 (89.3) | 297 (90.3) | 97 (86.6) | REF | - |
| Yes | 19 (4.3) | 13 (4.0) | 6 (5.4) | 1.41 [0.52-3.82] | - |
| Unsure | 28 (6.3) | 19 (5.8) | 9 (8.0) | 1.45 [0.64-3.31] | - |
| **Self-Reported HIV Status** |  |  |  |  |  |
| Positive | 0 (0) | 0 (0) | 0 (0) | - | - |
| **Total** | **441** | **329** | **112** |  |  |
| Negative | 108 (24.5) | 86 (26.1) | 22 (19.6) | REF | - |
| Unsure | 333 (75.5) | 243 (73.9) | 90 (80.4) | 1.45 [0.85-2.45] | - |
| **Self-Reported Pregnancy Status Among Females** |  |  |  |  |  |
| No | 254 (97.3) | 204 (96.7) | 50 (100.0) | - | - |
| Yes | 7 (2.7) | 7 (3.3) | 0 (0) | - | - |
| **Lived with someone who had TB for at least one day** |  |  |  |  |  |
| No | 376 (85.3) | 286 (86.9) | 90 (80.4) | REF | - |
| Yes | 59 (13.4) | 38 (11.6) | 21 (18.8) | 1.76 [0.98-3.15] | - |
| Unsure | 6 (1.4) | 5 (1.5) | 1 (0.9) | 0.64 [0.07-5.51] | - |

## DISCUSSION

In this study among HCWs in four healthcare facilities in Yogyakarta, no HCWs were diagnosed microbiologically or clinically with active TB disease, although 9% reported previous or current TB disease. TBI was diagnosed in 25% of those eligible for TST and who had the TST read; this was highest in staff from the hospital (31%, 95% CI: 26-37). A significant association was found between TBI and being male, currently working in the participating hospital and older age.

Annual incidence of active TB disease among HCWs has been reported to range from 69-5,780 per 100,000 population, and has consistently been documented to be higher than the general population (5). No active disease was diagnosed during the study, which could suggest low disease incidence in this population of HCWs, adequate disease diagnosis and detection programs within these health facilities, or could be due to the cross-sectional nature of the screening. However, self-reported past TB disease prevalence of 9% suggests disease incidence may be relatively high. This prevalence was similar to reported past disease among HCWs in a tertiary hospital in Bandung, Indonesia (11).

Prevalence of TBI (using TST) among HCWs in LMICs has been reported to range from 14–98% (mean 49%), and is higher among HCWs in high burden settings (pooled estimate of 55%, 95% CI 41-69%) (6). Our study findings of 25% were lower than this, and lower than a prevalence study conducted in a public tertiary hospital in Bandung, Indonesia that reported a 77% TST positivity (10,11). Our results, however, are similar to TBI prevalence studies across PHCs in Semarang, Indonesia (24%). Modelled community level TBI prevalence of 46% across Indonesia is also higher than the prevalence reported in our study (19). However, Yogyakarta province does have a lower estimated TB incidence (between 250-300 per 100,000 population) compared to the Indonesian average, and infection may parallel this trend, although prevalence estimates for TBI do not exist at this jurisdictional level (12).

Being male and increasing age as non-occupational risk factors for TBI among HCWs are consistent with several international studies (6,20–26). Male sex as a risk factor for TBI and disease is well documented (27,28), however the role of biological, social and structural factors for this increased risk is still undetermined, as is exposure risk behaviour (27). Other studies have found other non-occupational factors associated with TBI such as education level, smoking status, diabetes mellitus and household contact with a person with TB (5,6). Our study did not find an independent association between these factors and TBI in this population of HCWs.

The evidence is varied surrounding occupational factors as risk factors for TBI, such as years of work in the healthcare sector, contact with a TB patient, occupation type or work location (5,6,20,22,23,29). Our study did not find any individual occupational risk factors associated with TBI in health services that provide TB services, although we did find a significantly higher odds of infection among those working in the participating private hospital compared to the primary care facilities. This may allude to differences in provision of care, especially TB care, health facility IPC policy and procedure, and health facility infrastructure and layout. Persistent occupational TB exposure and poor levels of TBI control implementation in LMICs is a known significant factor for the increased risk of HCWs contracting TB (30). Higher risk of acquiring TB has been associated with certain work locations such as inpatient TB services, general medicine wards, emergency rooms and laboratories compared to medium risk for outpatient and surgical services, compared to the general populations (6). Similarly, certain occupations have had higher incidence of infection such as allied health, physicians and nurses (5,6).

Control measures may decrease annual TB incidence among HCWs by as much as 81% in high incidence settings (30). Even administrative controls alone, such as routine patient and HCW screening, have proven effective in decreasing the risk for TBI among HCWs in LMICs, suggesting these should be prioritised (5). The lack of implementation and scale-up of preventive measures, particularly failure to rapidly diagnose and treat TB, are primarily responsible for nosocomial transmission (30). The World Health Organization’s policy on TBI control in healthcare facilities recommends conducting routine screening and surveillance of TB disease among HCWs (4). Despite the benefits and recommendations, the implementation of these policies, IPC procedures and surveillance in high burden and/or LMICs is low (30). During the study period healthcare facilities in Indonesia did not routinely screen for active TB or TBI or provide preventive therapy for their staff, even though HCWs have been listed as high-risk in the 2016-2020 National Strategic Plan and should be provided with these services (31). Budget distribution, the lack of a national guideline for HCW screening and lack of healthcare facility awareness have been listed as reasons for this lack of implementation.

This study has some limitations. Primarily, the cross-sectional design and target population of all staff at pre-selected study sites meant that while all facilities in the sub-districts participated, the prevalence of TB among HCWs and characteristics associated with TBI may not be generalisable to the district, provincial or sub-national level. Ascertaining the acquisition source for infection was not possible, with community sources possible in addition to healthcare facility sources. Secondly, 14% of HCW registered at the facilities did not participate, close to 20% of eligible participants did not have a TST administered, and 10% were loss to follow up for TST reading. Attrition at each stage may have introduced bias and potentially over or under-estimated the prevalence and association estimates. The main reason for not having a TST administered was due to the recommendation to delay TST for 4-weeks post COVID-19 vaccination (32). The screening service did return after the vaccination roll-out at the facility, however, few participants returned. Thirdly, the limitations with TST’s sensitivity and specificity may have over or under estimated the prevalence of TBI (33). Fourthly, categorising HCW occupation, due to small numbers, may have skewed the association with TBI. Finally, besides BMI and TB investigations, all clinical characteristics were self-reported, which may have introduced bias, over or under-estimating the association between characteristics and TBI. Strengths of this study include high participation proportions across the facilities, the use of radiological and microbiological diagnostic tools, the collection of detailed demographic, occupational and clinical characteristics to investigate risk factors for TBI, and the participation of non-medical healthcare workers to explore occupational risk of TB.

## CONCLUSION

Indonesia is a high incidence setting for TB, and nosocomial and occupational TB hinder TB elimination efforts. This study supports prioritisation of HCWs as a high-risk group for TB infection and disease, and the need for comprehensive prevention and control programs in Indonesia. These include annual staff TB screening, ensuring implementation of IPC procedures in facilities, TB infection and disease surveillance, access to preventive therapy and clear national guidelines. Further, it identifies characteristics of HCWs in Yogyakarta at higher risk, identifying certain groups who could be prioritised in screening programs if universal coverage of prevention and control measures cannot be achieved.

## Data Availability

Data cannot be shared publicly because of project partnership stipulations/agreements. Data are available from the University of Gadjah Mada and Burnet Institute for researchers who meet the criteria for access to confidential data.

## ACKNOWLEDGEMENTS

The authors wish to thank the Zero TB Yogyakarta Programme, Yogyakarta Provincial and District Health offices, and the healthcare facilities and health care workers that participated; and Associate Professor, A Richardson, for her statistical expertise, review of the data analysis and subsequent suggestions.

The study was endorsed by the Indonesian NTP, Yogyakarta Provincial Government and Health Office, Yogyakarta City and Kulon Progo District Health Offices and associated health facilities involved in the research.

